# Outcomes of tracing antiretroviral therapy clients after missed clinic visits in South Africa: A retrospective cohort analysis

**DOI:** 10.64898/2026.11.24.26368899

**Authors:** N Mutanda, V Ntjikelane, E Kachingwe, L Sande, M Maskew, M Benade, M Manganye, L Malala, AR Marri, S Rosen

## Abstract

**Background:** In South Africa, interruption of antiretroviral therapy (ART) (missing a clinic visit or medication refill) for more than 28 days is very common, with an estimated 60% of clients interrupting at least once during their first year after initiation. Healthcare worker tracing of those who miss visits or medication refills, either through phone calls, text messages, or home visits (physical tracing), is a globally recommended, near-universal component of national HIV programs, but little is known about its effectiveness in returning clients to care.

**Methods:** We conducted a retrospective analysis of clients eligible for tracing under South Africa’s national guidelines (≥7 days late for a scheduled visit) at 15 public-sector ART clinics between January 2024 and August 2025. Tracing records were abstracted from facility paper registers and linked to electronic medical records to determine outcomes. We report tracing contact status (reached directly, reached indirectly through family member or friend, not reached, unknown) and final outcomes (return to care, self-return, no return, transfer/death, misidentification), separately for telephone and physical tracing, including time to return to care using Kaplan-Meier analysis.

**Results:** Tracing was attempted for 5,161 clients by telephone and 3,902 by physical visit. Roughly half of clients could be reached by tracers, either directly (43% telephone, 47% physical) or indirectly (7% telephone, 7% physical). Among those reached directly, 62-65% returned to care after contact. Among those who could not be reached, most nonetheless self-returned to care without any tracing contact (73% telephone, 65% physical), though more slowly than those who were reached. By day 90, two thirds of those reached directly by phone (68%) or physically (70%) had returned to care, compared to 60% and 53% of those not reached. Roughly 15% of tracing attempts targeted clients who had already returned to care, transferred, died, or had never actually interrupted care, reflecting data-quality challenges in the tracing and medical record systems.

**Conclusions:** Tracing HIV treatment clients who have missed clinic visits or medication pickups in South Africa appears to accelerate return to care but has an unclear effect on the overall proportion of those who return. Current data systems hamper efficient tracing efforts. Digitizing paper tracing registers, more targeted tracing that focuses limited resources on clients who genuinely require outreach, and interventions that promote client-initiated updates and two-way communication could allow better allocation of tracing resources to those truly in need.

## Introduction

As high-HIV prevalence countries in sub-Saharan Africa have approached global targets for epidemic control[1], retaining HIV-positive individuals on lifelong antiretroviral therapy (ART) has emerged as one of the main challenges facing national HIV treatment programs[2]. This is particularly true in South Africa, where a majority of clients who present at public sector clinics for treatment “initiation” are in fact non-naïve re-initiators who are returning after an interruption[3,4]. A recent analysis of routinely collected medical record data estimated disengagement from care at 23% by the end of their first year on ART[5], while another study reported that some 60% of all clients interrupt care at least once during their years on treatment[6].

A widely used intervention to address these high interruption rates is client tracing, a standard recommendation of the World Health Organization and included in most countries’ national ART guidelines[7]. Tracing (or tracking) uses health care workers, lay personnel, and/or mHealth technologies to contact clients who have missed a clinic visit or medication refill, with the intention of returning them to continuous care. Contact can be electronic (phone calls, SMS, or WhatsApp messages) or physical (home visits). Under current South African guidelines for tracing and recall[8], ART clients who are ≥7 days late for scheduled clinic visits or medication pickups should be traced, first by phone and then through home visits, with tracing priority assigned based on the client’s clinical risk (e.g. those with AHD disease or abnormal lab results are prioritized)[8]. Tracing is typically embedded within a package of other interventions to increase retention in care, such as differentiated service delivery models and multimonth dispensing of medications. In addition to encouraging timely return to care, tracing can help ascertain outcomes that would otherwise appear as disengagement, such as transfer to another facility and mortality.

Despite the near-universality of tracing as an intervention, there is very little known about its effectiveness in increasing retention in care in routine practice. Most estimates of the proportion of clients who return to care after tracing are based on non-routine study interventions, where external resources support intensive tracing efforts and documentation of results[7]. The duration of interruption required to trigger tracing, the timing of tracing after the interruption, the types and intensity of tracing efforts, and the interval during which return to care is attributed to tracing vary widely in the published literature, with no consistent indicators or reporting formats[7]. Existing data in South Africa suggest that a large proportion of clients who are late for scheduled visits return to care within one month of a missed interaction, with most returning within two weeks [5,9]. We cannot determine, however, whether these clients returned in response to tracing efforts or self-returned to care without any intervention, as tracing interactions are not captured in the national electronic medical record, TIER.Net. We have not found any estimates of the effectiveness of tracing in routine practice in increasing the proportion of clients who return to care after an interruption, nor is there any evidence available about how tracing is implemented in practice at facility level. Without this information, it is impossible to determine whether tracing is a worthwhile intervention or how to optimize its effectiveness.

Retaining clients in continuous care is essential to achieving HIV treatment goals[10], and tracing after a missed visit remains a core strategy to improve retention, despite its mixed results. To generate evidence to make tracing more efficient, we conducted a retrospective analysis of clients eligible for tracing under South Africa’s national guidelines between January 2024 and August 2025, extracting tracing records from paper registers and matching them to clients’ clinic records to determine outcomes. Here we report the primary outcome of return to care to the originating facility after tracing eligibility, stratified by whether or not tracing contact was made, and related results.

## Methods

We conducted a retrospective review of clinic tracing registers, which documented tracing activity by the clinic tracing teams, and linked register data to individuals’ electronic medical records to determine outcomes after tracing.

### Tracing guidelines and procedures in South Africa

Tracing guidelines for HIV treatment are described in South Africa’s Differentiated Models of Care Standard Operating Procedures, SOP 7 (Supplementary File 1), which specifies procedures for “Tracing and Recall” [8]. Facilities are advised to trace all chronic disease clients, including those on ART for HIV, who are late for a scheduled clinic or medication pickup visit by 7 days or more or for whom abnormal lab results have been received. Consent for SMS, phone call, and/or home visit tracing is obtained at a client’s first HIV-related clinic visit. Tracing attempts should first be by telephone (up to three calls), followed by home visits if calls are unsuccessful. If the tracer interacts with a family member or friend who answers the client’s phone or is present at the client’s home, a message may be left or a status of deceased recorded, but tracers. Tracers are not permitted to discuss a client’s status with a family member or friend, beyond being informed of a death.

Clients eligible for tracing are identified by the electronic medical record system, TIER.Net[11,12] which generates an “integrated missed appointment and unconfirmed lost to follow-up list” that includes all clients who have missed their scheduled return dates. A client’s details remain on the list until one of the following is confirmed: a clinic visit, death, transfer to another facility, or loss to follow-up, which is defined as an interruption >90 days. Facilities that retrieve client files from filing rooms daily, ahead of scheduled visits, also use the files remaining unclaimed at the end of the day to identify clients who need to be traced. Guidelines call for all tracing activities and client outcomes to be documented in the tracing register, the client’s file, and TIER.Net.

### Study sites and population

This study was conducted at 15 public sector clinics within the previously described SENTINEL South Africa network[13–16]. Study sites were located in Ehlanzeni District in Mpumalanga Province (5 sites), King Cetshwayo District in KwaZulu-Natal Province (6 sites), and West Rand District in Gauteng Province (4 sites). The selected facilities were all public sector primary healthcare clinics with large ART client volumes and reflected a mix of rural and urban settings.

All clients aged 18 or older who were receiving ART at the study sites and for whom tracing was attempted between January 1, 2024 and August 4, 2025 and recorded in the tracing registers were eligible for the study. Where clients missed multiple visits during the study period, only the first was analyzed.

### Datasets

The study used two separate datasets for this analysis. First, the study team constructed a dataset from paper registers for telephone and physical tracing at the study sites. In a pre-study assessment of tracing procedures at the study sites, we found a total of ten different types of tracing documentation [17]. The primary source for keeping track of tracing activities is facility tracing registers, into which data from the TIER.Net missed visits lists mentioned above are transcribed. Facilities indicated that tracing data may also be recorded in community health workers’ tracing registers, referral forms, and individual client paper files, however[17]. At each study site, research assistants identified all available facility-level tracing registers from the study observation period (January 1, 2024 to August 4, 2025). They extracted individual client data and captured it in a REDCap database between June 3, 2025 and October 6, 2025.

The second dataset comprised routinely generated medical record data extracted from TIER.Net for all ART clients in the three districts. The observation period started on 1 January 2024 in each district and ended on 6 November 2025 in West Rand, 6 January 2026 in Ehlanzeni, and 7 January 2026 in King Cetshwayo, reflecting the date of the last recorded entry in each district at the time of data extraction. The median follow-up time was 8 months (IQR 6,12) in West Rand and 15 months (IQR 12,18) in King Cetshwayo for telephonic tracing and 11 months (IQR 7,16) in Ehlanzeni and 16 months (IQR 12,19) in West Rand for physical tracing (Table 2). The minimum follow-up time after entry in the tracing register was 3 months in West Rand and 5 months in both Ehlanzeni and King Cetshwayo. This system captures scheduled visit dates, actual visit attendance dates, and client outcomes such as transfers, death, loss to follow-up, and retention in care for those attending their visits at the facility. The first and second datasets were then linked using client identifiers (name, surname, patient file, date of birth, ART start date, and gender) to assess tracing outcomes for each client.

TIER.Net data were accessed for research purposes on 8 January 2026 for Ehlanzeni District, 8 January 2026 for King Cetshwayo District, and 25 November 2025 for West Rand District.

### Study outcomes

The primary outcome for the analysis reported here is return to care at any time in the observation period (yes/no) among clients eligible for tracing under national guidelines. The exposure is the result of a recorded tracing attempt (reached/not reached/unknown). We compared the proportion who returned to care between clients who were reached during a tracing attempt and clients who could not be reached. Among those reached and those who could not be reached, we also describe the proportion who did not return to care. We first assigned tracing contact status based on whether the client was successfully reached during tracing, with three categories: reached, not reached, and unknown (Table 1, upper half). Using the linked data, we then further classified each client’s final tracing outcome, i.e. whether they returned to care following the tracing contact among those reached during the tracing attempt, self-returned to care among those not reached during tracing, or did not return. To do this, we used the date of the missed visit that triggered tracing, the date of tracing, and any subsequent recorded visit (return date) after the missed date, indicating that the client had returned to care (Table 1, lower half). For all clients who returned to care at any time during the observation period, we calculated the time taken to return in days, defined as the number of days between the missed visit and the first visit after the missed visit. (Note that “return to care” refers to the originating clinic; clients who informally transfer to a different facility will be reported as not returned.)

**Table 1.** Study metrics and outcomes.

| **Metric** | **Definition** |
|---|---|
| ***Tracing contact categories*** |  |
| Reached-directly | Reached client at any time during the follow up period. Where the client was reached directly, the result of the contact was a) a promise to return on a specific date, b) a reported transfer to another facility, or c) a statement that the client is unwilling to return to care. |
| Reached-indirectly | Where a family member or friend was reached instead, the result of the contact was recorded as a) message for the client left with a relative or other available person, or b) the client was reported as deceased. |
| Not reached | Could not reach client, family member, or friend. For telephonic tracing, included voicemail, invalid number, wrong number, or unanswered calls. For physical tracing, included unknown at the address, migrated or relocated, wrong address, or a locked gate. |
| Unknown | No tracing result recorded in any data source; unable to determine if client was reached. |
| ***Final tracing outcomes*** |  |
| Reached and returned after tracing contact | Reached during tracing; visit was recorded after the date of the tracing contact. |
| Not reached by tracing but self-returned | Not reached during a tracing attempt ; visit was recorded after the missed visit. |
| Self-returned before tracing attempt | Client had already returned to care before the tracing attempt but remained on the tracing list. |
| Did not return to care at the site (reached or not reached) | No visit was recorded after the missed visit. |
| Transferred or died | Outcomes recorded in facility registers and/or reported during tracing attempt. |
| Misclassified as eligible for tracing (reached or not reached) | Client who appeared on the tracing list due to data errors or delays in data capture, despite not having missed a visit. |

To illustrate the order and timing of tracing attempts and outcomes, we also developed tracing flow charts, as shown in Figure 1. The first row (Cell 1) represents clients for whom a documented tracing attempt was made for a client in the study population. The second row of Figure 1 (Cells 2a, 2b, 2c) captures tracing attempts and indicates whether or not the client or a family member or friend was reached. Box 2c includes those for whom registers show that an attempt was made but not whether anyone was reached. Finally, the last rows of Figure 1 (Cells 3a-e, 4a-f, 5a-c and 6a-c) report the outcomes of those for whom tracing was attempted. This last row is intended to be both comprehensive and mutually exclusive, so that every ART client for whom at least one tracing attempt was made and documented can be assigned a final outcome box. Cell 3a represents the primary outcome for our study, return to care after a tracing contact. With observational data, it is not possible to determine whether the tracing contact directly caused the return to care, but the proportion of tracing-eligible clients who end up in Cell 3a is typically regarded as the success rate of a tracing intervention. (For simplicity, the last row of Figure 1 does not specify time to return to care; this information will be reported in a separate figure.)

**Figure 1.**
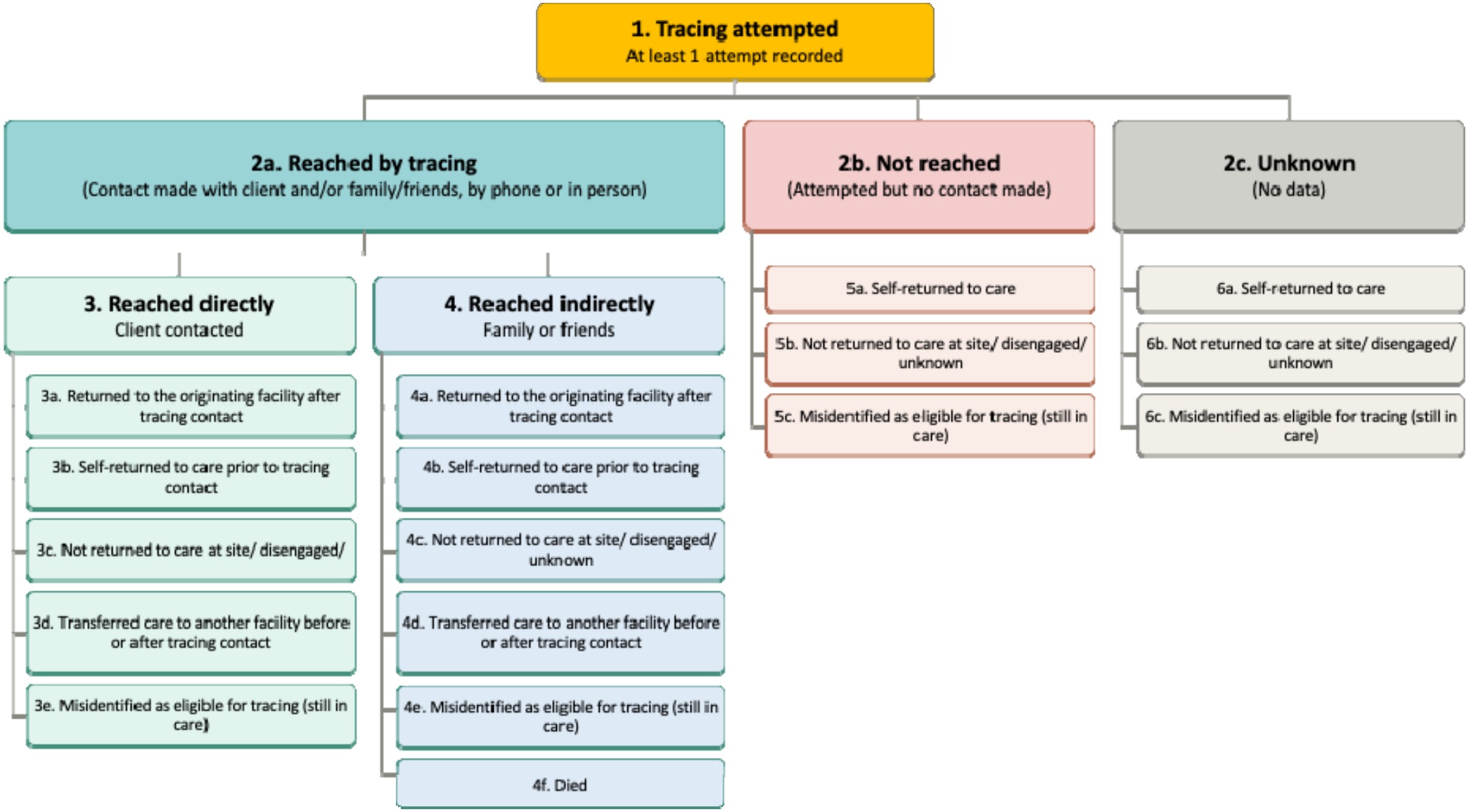
Study outcomes

### Data analysis

Results are presented separately for each tracing method (telephone and physical tracing) to assess whether the proportion successfully contacted and returned to care varied by method. For West Rand District, where data were available for both tracing methods, each tracing attempt was treated separately under its respective method. Clients who were traced by both methods therefore contributed one attempt to each method and appear in both sets of results. We report the number of clients traced by both methods separately. Tracing contact and outcomes were described using frequencies and proportions for categorical variables and medians with interquartile ranges for continuous variables.

We note that although guidelines recommend up to three telephone tracing attempts followed by up to three home visits, actual tracing practices varied across sites in number and sequence, depending on available resources and site preferences. Due to the large number of missing registers, we could not track individuals longitudinally across telephone and physical tracing attempts. Each tracing method was therefore analyzed separately.

Among clients who returned to care, the time to return following a missed clinic visit was estimated using Kaplan-Meier survival analysis. For each client, the event of interest was return to care, defined as the first clinic visit occurring after a missed scheduled visit. Person time accrued from date of missed scheduled visit to the earliest of: 1) date of return visit; 2) death; 3) transfer out; 4) database censor date; 5) 90 days follow up. Clients who had not returned to care within 90 days were censored.

Kaplan-Meier curves were generated separately for telephone tracing and physical tracing, stratified by tracing contact outcome: (1) reached directly during tracing, (2) reached indirectly during tracing, (3) not reached during tracing, and (4) tracing contact unknown. This analysis was limited to the first 90 days after each missed visit. Each curve depicts the cumulative incidence plot (1 − survival) of the probability of returning to care over 90 days after a missed visit, by tracing contact outcome allowing visual comparison of the pace of return to care across groups. Differences in survival distributions between groups were assessed using the log-rank test, with a two-sided p-value <0.05 considered statistically significant. The Kaplan-Meier curves were plotted to illustrate differences in return-to-care patterns by tracing contact outcome for both telephone and physical tracing methods.

### Ethics approval

The use of data for this analysis was approved by the University of the Witwatersrand Human Research Ethics Committee (M2409113). De-identified data were shared with Boston University and approved by the Boston University Institutional Review Board (H-38115). The protocol was also approved by the respective District and Provincial Health Research Committees through the National Health Research Database for each study district. We received a waiver of informed consent for all study data.

### Artificial intelligence disclosure statement

ChatGPT-6 Luna and Claude Sonnet 5.5 were utilized during the preparation of this manuscript to design figures from data provided, check internal consistency of data and results reported, edit text for clarity, and copyedit text. The authors collected and analyzed all data, reviewed all AI contributions, and confirmed all citations. The authors take full responsibility for the accuracy, integrity, and final content of this publication.

## Results

### Tracing data

Research staff were able to locate telephone and/or physical tracing registers covering anywhere from 0 to 18 months of the 20-month study period, from January 1 2024 to August 4 2025. Figure 2 indicates the months for which registers were available, by study site.

**Figure 2.**
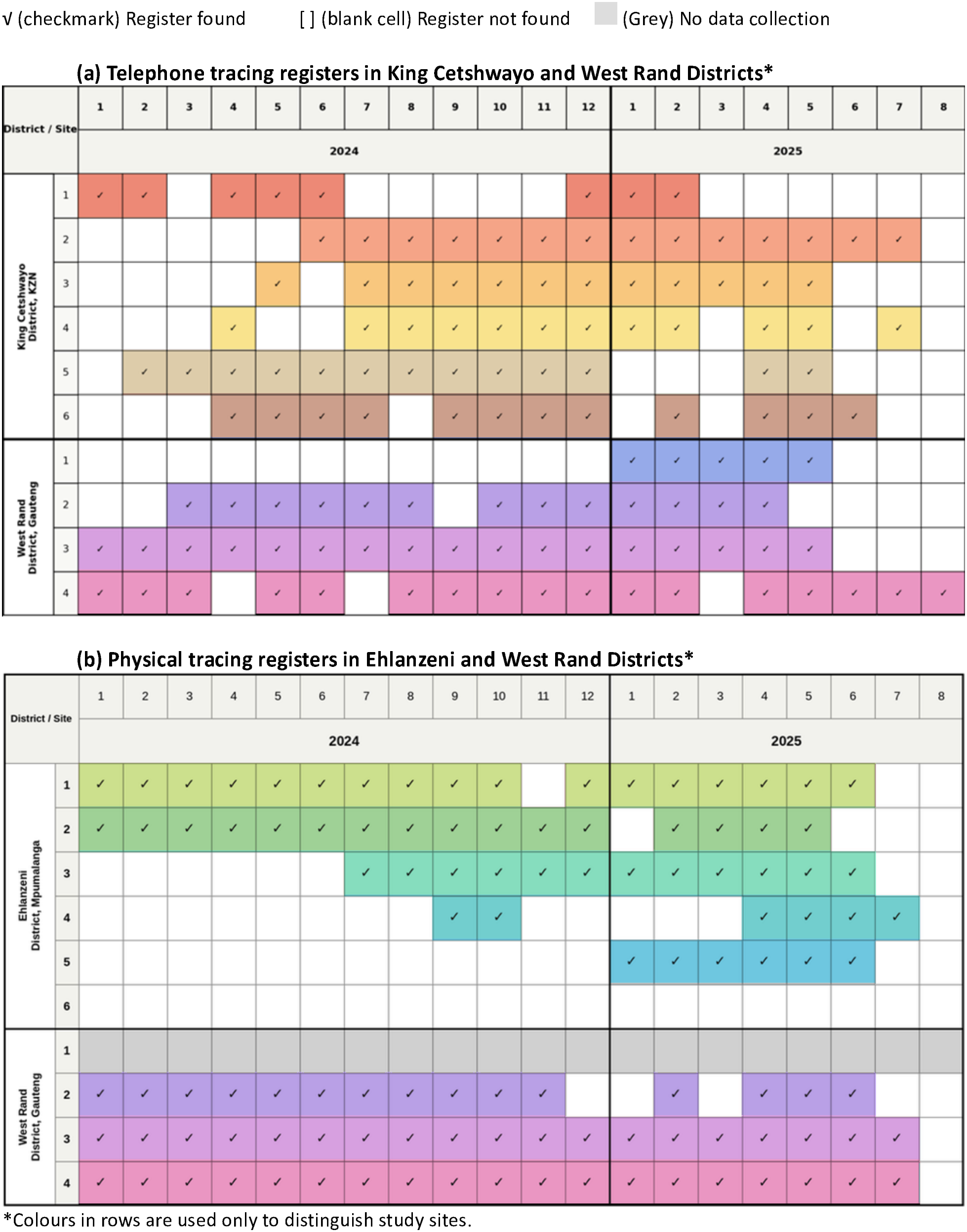
Telephone (a) and physical (b) tracing registers available at study sites*

All facilities in KC and WR districts utilized standard tracing registers issued by the Department of Health to document tracing activities. Sites in KC were able to provide telephone tracing registers covering between 8 and 14 months of the 20-month study period. We found only a very small number of physical tracing records for the KC sites during the study period and excluded them from analysis due to the large number of missing registers. In WR, three of the four sites included had fairly complete sets of telephone registers available, covering between 13 and 17 months of the 20-month period. The fourth site (West Rand Site 1 in Figure 2a) had no registers on hand for 2024. Physical tracing analysis for West Rand sites was restricted to 3 facilities; limited study resources prevented entry of physical tracing register data for the fourth site.

In Ehlanzeni District, facilities used different types of registers to record tracing activities, mainly because tracing was carried out by implementing partner organizations which utilized their own register formats. Of the six study facilities in EH, one had no telephone or physical tracing registers for the study period available at all (Figure 2b Ehlanzeni Site 6). This site indicated that phone contact for tracing may be recorded directly into client files or not recorded. The remaining five facilities could provide physical tracing registers only. Some facility managers explained that telephone tracing registers had been temporarily removed by former implementing partners following PEPFAR funding termination and had not yet been returned at the time of study data collection.

### Study sample

Numbers of clients enrolled in the study, by type of tracing attempted and district, are shown in Table 2. Detailed flow charts describing the construction of each district’s sample are provided in Supplementary Figures 1-4. The numbers enrolled reflected multiple variables, including the availability of tracing registers (Figure 2), each site’s diligence in documenting tracing attempts, and the number of clients for whom tracing was attempted, which in turn reflects both provider and client behaviour. As Table 2 indicates, client characteristics were broadly similar across districts and tracing types. The majority of traced clients (86-91%) had been on ART for more than a year. Intensity of tracing effort (number of times tracing was attempted) varied somewhat across districts and type of tracing; however, in all districts, most clients were traced only once.

**Table 2.**
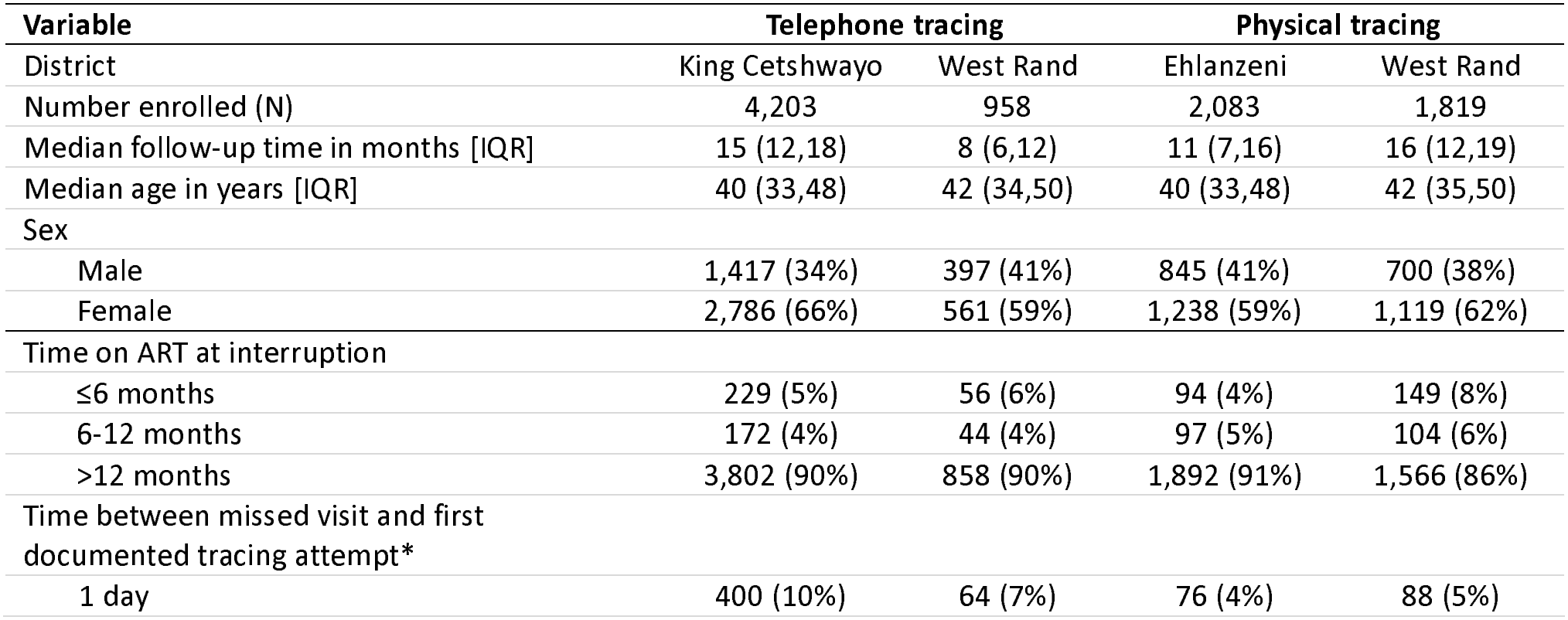
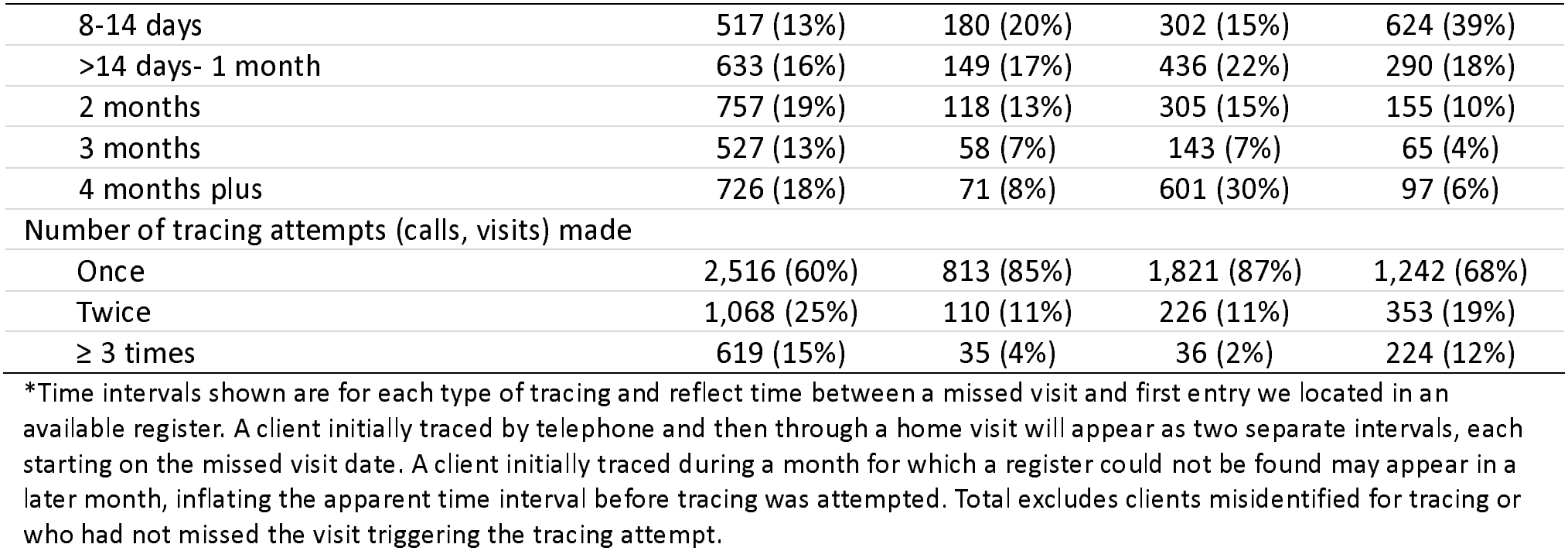
Clients enrolled in the study.

### Tracing outcomes

Outcomes of tracing attempts for each type of tracing are illustrated in Figure 3 and reported in Table3. For both telephone and physical tracing, roughly half of those called or visited could be reached either directly or indirectly, averaging 49% for telephone tracing and 54% for physical tracing. Of those reached, contact was direct for 86% and indirect (message left with family or friend) for 14% with telephone tracing and direct for 87% and indirect for 13% with physical tracing. Among those who were successfully reached directly, 65% returned to the originating facility after telephone contact and 62% after physical tracing, with a tight range across districts and tracing types of just 61-67%. Of those reached indirectly, 67% returned to care after telephone tracing, while 66% returned following physical tracing. Among all clients for whom tracing was attempted, regardless of whether contact was made, 32% (1,653/5,161) returned after telephone tracing and 34% (1,324/3,902) after physical tracing.

**Figure 3:**
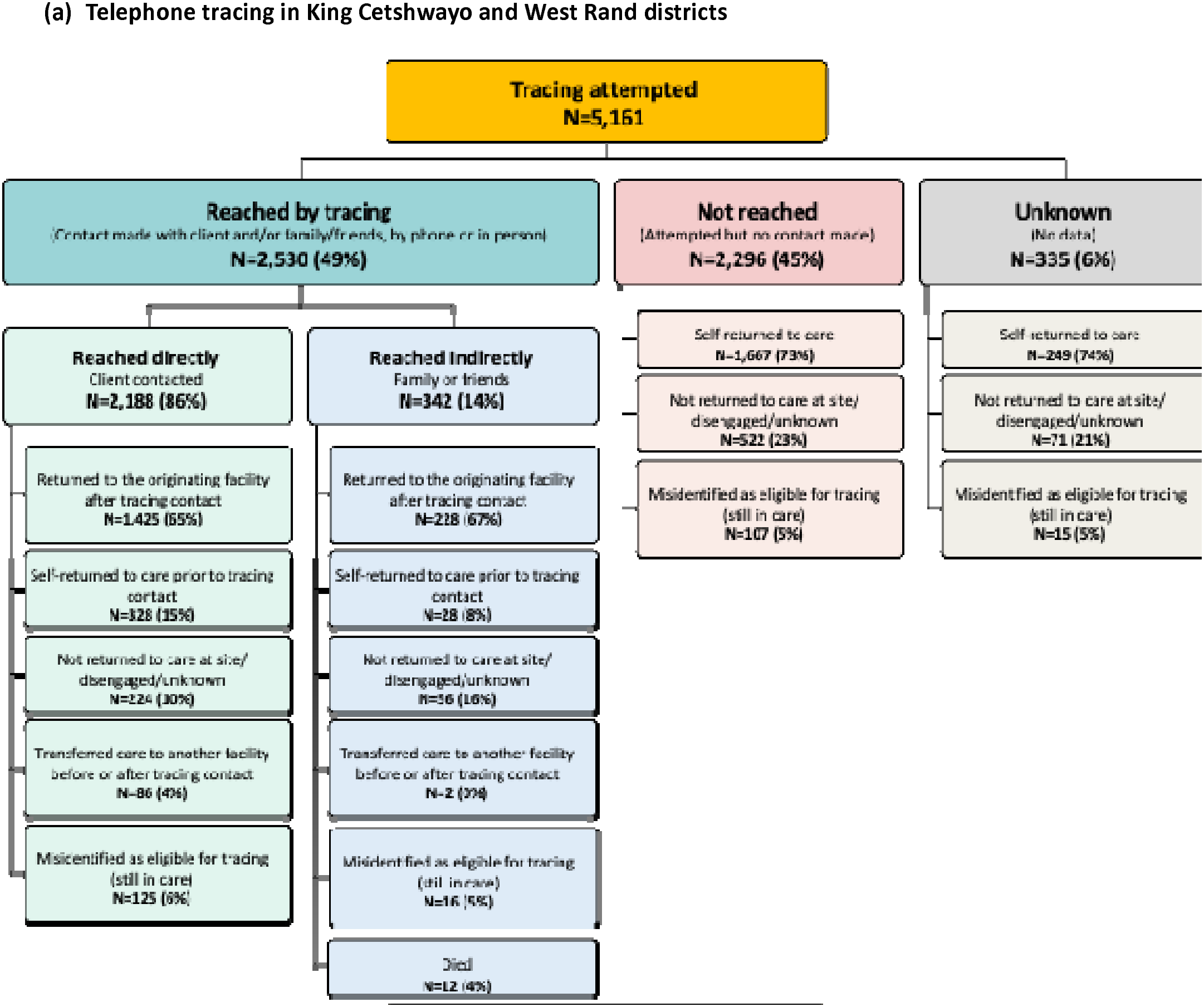
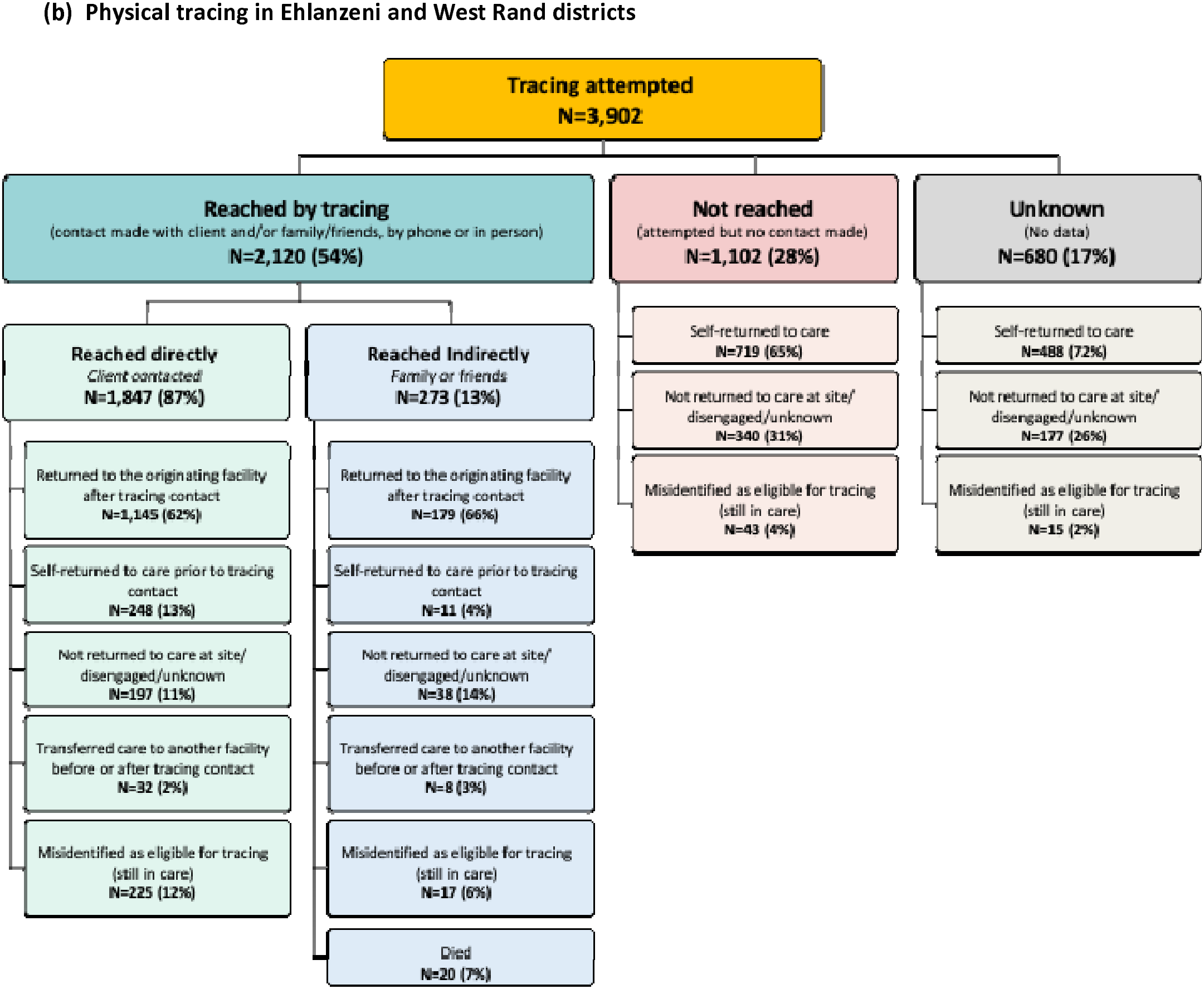
Tracing outcomes for (a) telephone tracing in King Cetshwayo and West Rand districts and (b) physical tracing in Ehlanzeni and West Rand districts

Among clients for whom tracing was attempted but who could not be reached, 73% of those who could not be reached by phone and 65% of those who could not be reached via physical tracing ultimately returned to care. A larger proportion of this group was classified as not returned—23% for telephone and 31% for physical tracing, compared to a range of 10-16% among clients who were reached either reached directly or indirectly using either approach.

Table 3 also indicates that 4-15% of those for whom tracing was attempted and who were reached either directly or indirectly by either method had already returned to care at the originating facility before the attempt was made (self-returned before tracing contact) and that 6-11% had never interrupted care at all, highlighting the challenge posed by inaccurate and/or delayed data entry. This issue was also noted among clients who were not reached (4-5%) and those whose outcome was unknown (2-5%): medical records showed that the flagged visit had not actually been missed or that a transfer had been documented before the next scheduled visit. Additional client characteristics by tracing outcome are reported in Supplementary Table 1.

**Table 3.** Tracing outcomes by type and district (a) telephone tracing (b) physical tracing.

| Outcome | King Cetshwayo<br>n (%) | West Rand<br>n (%) | Total<br>n (%) |
|---|---|---|---|
| Number of clients for whom telephone tracing was attempted | 4,203 (100%) | 958 (100%) | 5,161 (100%) |
| ***Reached by tracing*** | 2,095 (50%) | 435 (45%) | 2,530 (49%) |
| *Reached directly* | 1,802 (86%) | 386 (89%) | 2,188 (86%) |
| Returned to originating facility after tracing contact | 1,165 (65%) | 260 (67%) | 1,425 (65%) |
| Self-returned to care prior to tracing contact | 312 (17%) | 16 (4%) | 328 (15%) |
| Not returned to care site/ disengaged / outcome unknown | 170 (9%) | 54 (14%) | 224 (10%) |
| Transferred care to another facility before or after a tracing contact | 72 (4%) | 14 (4%) | 86 (4%) |
| Misidentified as eligible for tracing (still in care) | 83 (5%) | 42 (11%) | 125 (6%) |
| *Reached indirectly* | 293 (14%) | 49 (11%) | 342 (14%) |
| Returned to originating facility after tracing contact | 195 (67%) | 33 (67%) | 228 (67%) |
| Self-returned to care prior to tracing contact | 27 (9%) | 1 (2%) | 28 (8%) |
| Not returned to care site/ disengaged / outcome unknown | 45 (15%) | 11 (22%) | 56 (16%) |
| Transferred care to another facility before or after a tracing contact | 0 (0%) | 2 (4%) | 2 (0%) |
| Misidentified as eligible for tracing (still in care) | 15 (5%) | 1 (2%) | 16 (5%) |
| Died | 11 (4%) | 1 (2%) | 12 (4%) |
| ***Not reached by tracing*** | 1,847 (44%) | 449 (47%) | 2,296 (45%) |
| Self-returned to care | 1,360 (74%) | 307 (68%) | 1,667 (73%) |
| Not returned to care at site/ disengaged / outcome unknown | 396 (21%) | 126 (28%) | 522 (23%) |
| Misidentified as eligible for tracing | 91 (5%) | 16 (4%) | 107 (5%) |
| ***Unknown if reached (no tracing outcome recorded)*** | 261 (6%) | 74 (8%) | 335 (6%) |
| Self-returned to care | 202 (77%) | 47 (64%) | 249 (74%) |
| Not returned to care at site/ disengaged / outcome unknown | 48 (18%) | 23 (31%) | 71 (21%) |
| Misidentified as eligible for tracing | 11 (4%) | 4 (5%) | 15 (5%) |
| Outcome | Ehlanzeni<br>n (%) | West Rand<br>n (%) | Total<br>n (%) |
| Number of clients for whom physical tracing was attempted | 2,083 (100%) | 1,819 (100%) | 3,902 (100%) |
| ***Reached by tracing*** | 1,116 (54%) | 1,004 (55%) | 2,120 (54%) |
| *Reached directly* | 1,062 (95%) | 785 (78%) | 1,847 (87%) |
| Returned to originating facility after tracing contact | 646 (61%) | 499 (64%) | 1,145 (62%) |
| Self-returned to care prior to tracing contact | 185 (17%) | 63 (8%) | 248 (13%) |
| Not returned to care site / disengaged / outcome unknown | 112 (11%) | 85 (11%) | 197 (11%) |
| Transferred care to another site before or after tracing contact | 20 (2%) | 12 (2%) | 32 (2%) |
| Misidentified as eligible for tracing (still in care) | 99 (9%) | 126 (16%) | 225 (12%) |
| *Reached indirectly* | 54 (5%) | 219 (22%) | 273 (13%) |
| Returned to originating facility after tracing contact | 22 (41%) | 157 (71%) | 179 (66%) |
| Self-returned to care prior to tracing contact | 5 (9%) | 6 (3%) | 11 (4%) |
| Not returned to care site / disengaged / outcome unknown | 9 (17%) | 29 (13%) | 38 (14%) |
| Transferred care to another site before or after tracing contact | 3 (6%) | 5 (2%) | 8 (3%) |
| Misidentified as eligible for tracing (still in care) | 1 (2%) | 16 (7%) | 17 (6%) |
| Died | 14 (26%) | 6 (3%) | 20 (7%) |
| ***Not reached by tracing*** | 394 (19%) | 708 (39%) | 1,102 (28%) |
| Self-returned to care | 208 (53%) | 511 (72%) | 719 (65%) |
| Not returned to care site / disengaged / outcome unknown | 186 (47%) | 154 (22%) | 340 (31%) |
| Misidentified as eligible for tracing | 0 (0%) | 43 (6%) | 43 (4%) |
| ***Unknown if reached (no tracing outcome recorded)*** | 573 (28%) | 107 (6%) | 680 (17%) |
| Self-returned to care | 412 (72%) | 76 (71%) | 488 (72%) |
| Not returned to care site/ disengaged / outcome unknown | 161 (28%) | 16 (15%) | 177 (26%) |
| Misidentified as eligible for tracing | 0 (0%) | 15 (14%) | 15 (2%) |

### Time to return to care

Figures 4a and 4b show time to return to care after a missed visit for all those who did return, by tracing type and outcome. Among clients who self-returned to care before telephone or physical tracing occurred and were reached directly or indirectly, most did so within 30 days of the missed visit (71% - 100%). Among clients who returned to care after being reached directly by tracing, approximately half returned within 30 days (52% for telephone tracing and 58% for physical tracing), while a further 34% and 38% returned after 30 days but within 90 days. Return times were slightly longer among clients who were not reached during tracing, with 44% % self-returning to care within 30 days of the missed visit for telephone tracing and 46% for physical tracing, and 36% and 33%, respectively, returning within 90 days.

**Figure 4.**
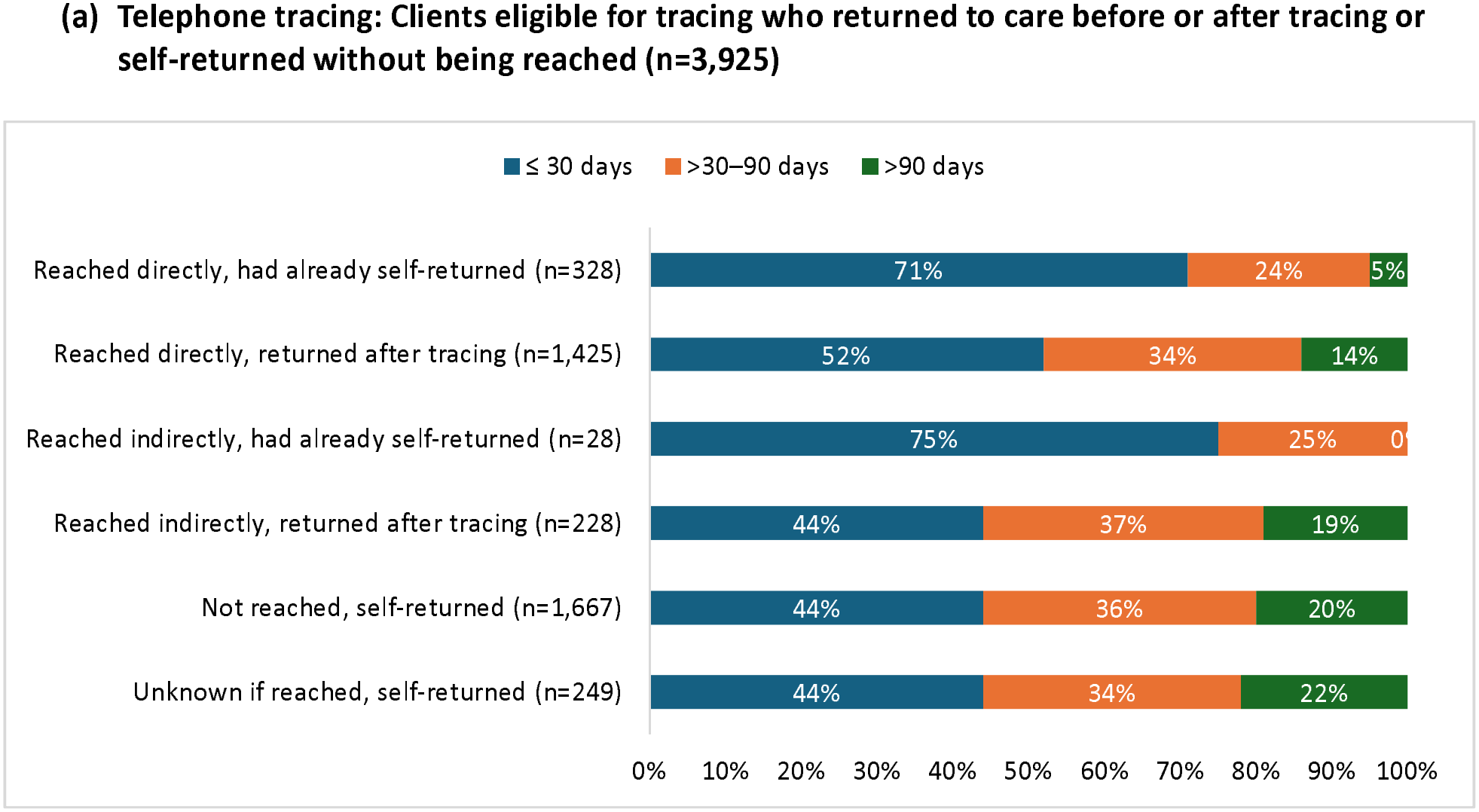
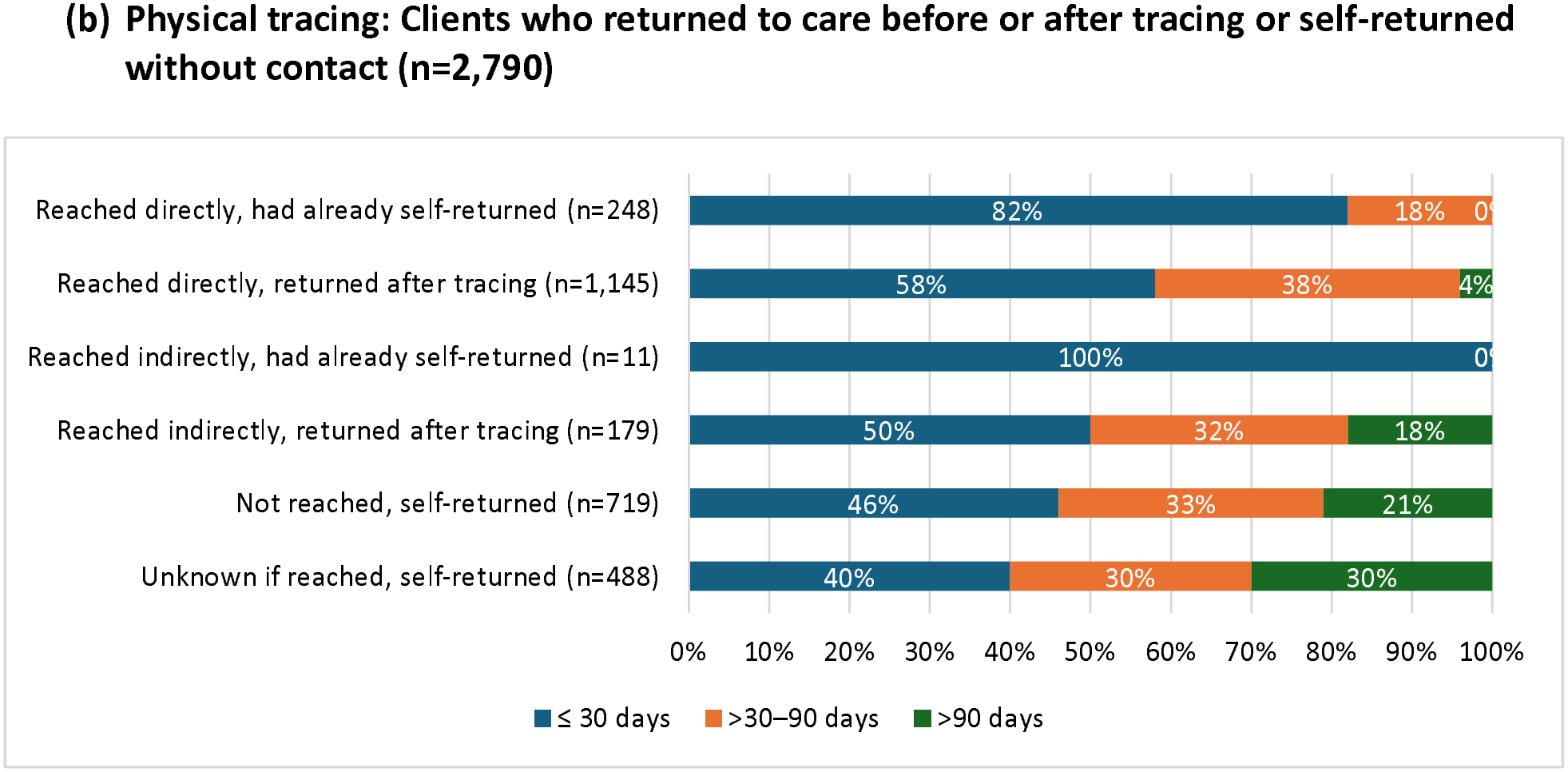
Time to return to care by outcome for (a) telephone tracing and (b) physical tracing

As shown in Figures 5a and 5b, the probability of returning to care after a missed visit differed by tracing outcome (reached directly, reached indirectly, not reached, unknown). Among clients traced by phone, 43% of those reached directly (green line) had returned to care by day 30, compared with 33% of those reached indirectly (blue line) and 33% of those not reached (red line). By day 90, 68% of those reached had returned, compared with 59% of those reached indirectly and 60% of those not reached. Among clients traced physically, almost 50% of those reached directly had returned by day 30, compared with 36% of those reached indirectly and 30% of those not reached, and by day 90 about 70% of those reached directly had returned, compared with 59% of those reached indirectly and 53% of those not reached. Notably, clients who were directly reached during tracing (green line) returned faster than all groups.

**Figure 5.**
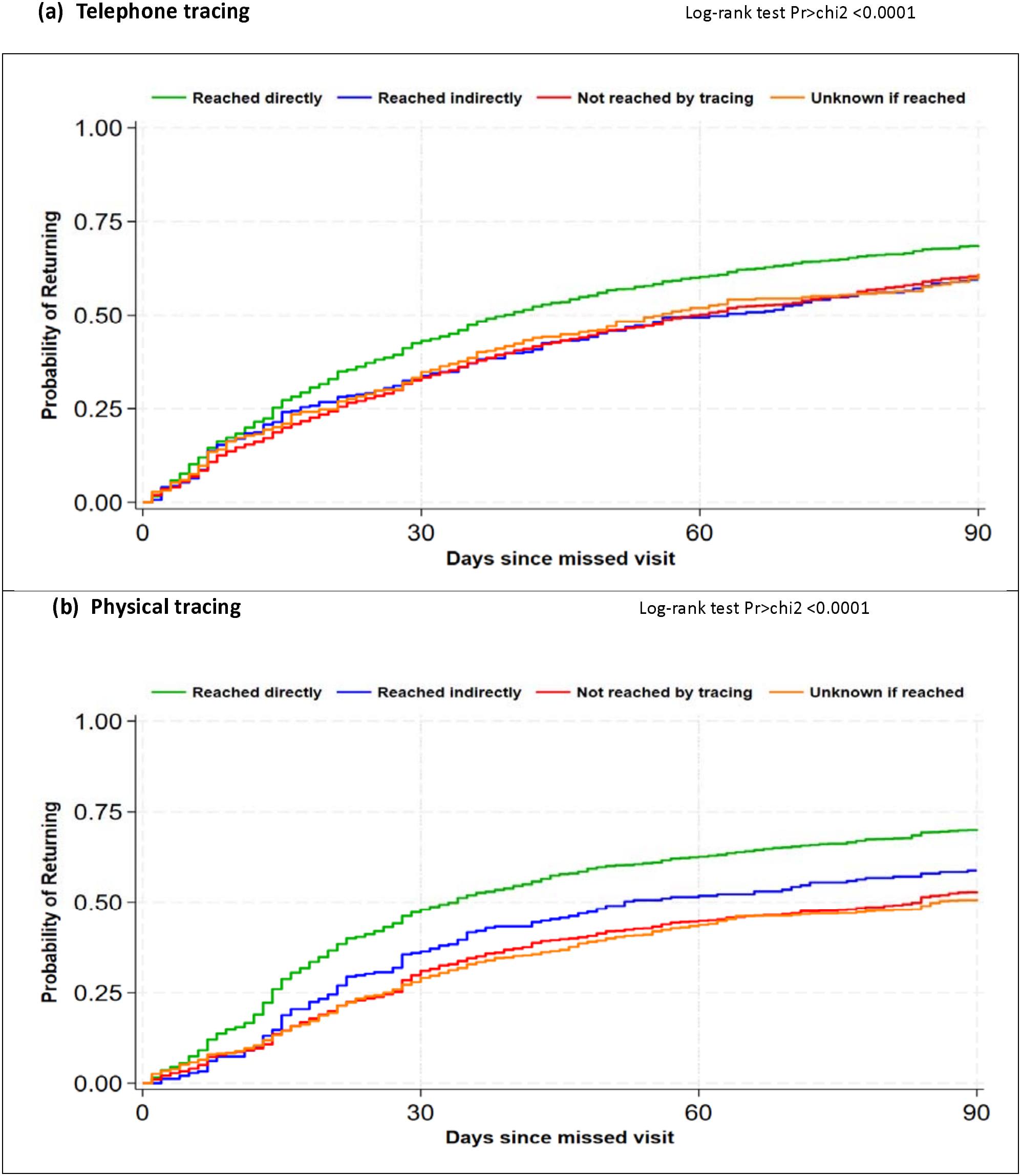
Probability of returning to care during the first 90 days after a missed visit, by tracing type and outcome (Kaplan-Meier analysis)

## Discussion

In this analysis of primary data from 15 public sector clinics in three districts in South Africa, we found that approximately half of the clients who interrupted HIV treatment and for whom tracing was attempted could be reached by either telephone or physical tracing. Of the 9,063 clients for whom either type of tracing was attempted, 33% (2,977) returned to care after successful tracing contact and 26% (2,386) returned without being reached during tracing.

Among those who could be reached directly, roughly two-thirds returned to care after tracing contact was made, and more than half of these returned within 30 days of missing a visit. Among those for whom tracing was attempted but who could not be reached, 70% (2,386/3,398) returned to care, although fewer of these self-returners came back within 30 days of missing a visit. In reporting these results, we note that individuals who can be reached by tracers may differ systematically from those who cannot and that the probability of returning to care after an interruption may be affected by other interventions beyond tracing. In addition, 14% of clients for whom tracing was attempted fell into categories for which tracing should not have been conducted at all: those who had already returned to care or transferred before the tracing attempt was made and those misclassified as eligible for tracing despite never having interrupted care. Unexpectedly, most clients who were traced were long-established on treatment: nearly 90% had been on ART for more than a year, while fewer than 10% had been on treatment for six months or less.

We believe that the results reported here suggest six important takeaways. First, and generally consistent with prior literature, only about half of the effort invested in tracing leads to actual contact with the target client [6]. In our study, this proportion did not differ widely by either province or tracing approach. Among clients who could not be reached, tracing registers documented several reasons for unsuccessful telephonic tracing, including missing contact details, invalid or incorrect numbers, voicemail, and unanswered calls. For physical tracing, documented reasons included clients being unknown at the recorded address, having moved or relocated, locked gates, and incorrect addresses. These challenges have been documented in the literature and help explain why tracing efforts often fail to reach all target clients [18–20].

A key contributor to the inability of tracers to reach target clients is the lack of accurate and up-to-date contact details in patient files. South African guidelines require clients to provide contact details and consent for their preferred tracing method at ART initiation. These details are expected to be reviewed and updated at subsequent clinic visits, but the consistency with which this is implemented is unclear. Consequently, contact information in patient records may become outdated over time. Other explanations may include clients intentionally providing incorrect contact information, reluctance to engage with tracers, changes in telephone numbers or residence, and provider clerical errors when transferring contact details from TIER.Net lists to tracing registers or notebooks. Fortunately, many of these problems can be solved, at least in part, through small adjustments to facility procedures.

Second, we found that tracing contact was followed by a return to care for approximately two-thirds of clients who could be reached. Using observational data, we cannot determine whether tracing itself increased the likelihood of return. Notably, substantial proportions of clients who could not be reached also returned to care: 73% of those not reached by phone and 65% of those not reached through physical tracing subsequently returned without any tracing contact. The value of tracing may be in accelerating these returns: return to care occurred more rapidly among those who were successfully reached directly, with 68% (telephone) and 70% (physical) back in care within 90 days, versus 60% and 53% among those not reached. The survival analysis similarly showed a higher probability of remaining out of care among those not reached compared to those reached during the first 90 days after a missed visit.

Our findings are consistent with existing evidence that most clients who miss scheduled visits do return to care within 90 days [5,8]. We note that a missed visit does not necessarily indicate treatment interruption, as clients may continue ART using leftover medication from previous dispensing, medication obtained from others, or medication purchased elsewhere. These findings raise an important question of who should be prioritized for tracing: a substantial share of eligible clients return on their own, and some of the fastest returners never needed to be traced at all. Prioritization should distinguish between clients at high risk of harm if they remain out of care and clients with a high probability of benefiting from tracing. The two groups may overlap, but they are not identical [7]. Profiling clients who miss appointments could help identify those at genuinely greater risk of prolonged disengagement and of harms due to interruptions, and support a more targeted approach to tracing rather than triggering tracing solely based on a missed appointment.

Third, the timing of tracing matters, but the optimal window for tracing remains unclear. Clients who returned before a tracing contact did so within a few days of the missed visit, with a median interval of 13 days (IQR 4,37) after the missed visit date for those for whom telephone tracing was attempted and 8 days (IQR 3,23) for those physically traced (supplementary Table 1). Tracing too soon—on the day after every missed visit--may therefore waste resources on clients who are already on their way back to care. Waiting too long, however, may reduce the opportunity to intervene before a short interruption becomes a prolonged one. The literature is inconsistent on this question, suggesting that further research on the optimal window for tracing after a missed visit is warranted.

Fourth, measuring the effectiveness of tracing is complicated by weak linkage between tracing records and electronic medical records (TIER.Net). Tracing activities are primarily recorded in paper-based registers, with the expectation that client outcomes will subsequently be captured in TIER.Net, but the two sources are not routinely reconciled. Discrepancies between the two sources abounded in our study. Clients labelled as lost to follow-up in tracing registers were sometimes shown in TIER.Net to have returned to and still be in care, based on recorded subsequent clinic visits. Delays in capturing patient files or updating visit dates in TIER.Net resulted in clients being incorrectly flagged for tracing. TIER.Net may also flag clients as having missed a visit when they attended the facility before their scheduled appointment date, as the system continues to identify the original scheduled date as missed. Although manually checking the client file before initiating tracing is recommended, this does not always occur, resulting in the tracing of clients who have already returned to care and further inefficient use of limited tracing resources.

In addition, because of the fragmented nature of the available documentation, we were unable to estimate the proportion of all clients who met tracing criteria (i.e. were >7 days late for a visit) whom we were able to identify and enroll in our study. The number enrolled in the study reflected the availability of registers for data extraction, the diligence with which tracing attempts were documented, the availability of staff to conduct tracing, each site’s own decisions about tracing priorities and timing, and the rate of treatment interruptions among each site’s client population. This challenge has also been reported in other South African studies, where discrepancies and misclassification of patient outcomes between tracing/outreach records and the national HIV treatment database, potentially distorting program forecasting, monitoring and evaluation, have been documented [18,21], and in multi-country tracing studies [19,22]. Integrated systems could improve data linkage, reduce inconsistencies across data sources, and generate timely, reliable information to support evidence-based decision-making.

Fifth, the study found that tracing data were difficult to locate and collect. At all 15 clinics, some tracing registers were missing, and most of those found were incomplete, lacking time periods, client data, tracing details, and/or outcomes. Lists of clients who had interrupted care were generated in TIER.Net and manually transcribed into paper registers, and tracing outcomes (whether the client was reached and returned to care) were recorded inconsistently. Etoori et al. 2020 [18] found similarly poor documentation of tracing activities and record-keeping in rural sites in South Africa.

Finally, we found that most clients for whom tracing was attempted in this study (90% overall) had already been on ART for more than a year at the time of the missed visit that triggered tracing. Since some 97% of all ART clients in South Africa had been on ART for more than one year as of 2025[23], this finding likely largely reflects the composition of the treatment cohort and thus the proportion all visits occurring after a year on ART. We note from Supplementary Table 1 that those on ART for less than 6 months were substantially less likely to return to care at all than were those with longer experience at the study sites, but we cannot determine whether this reflects a higher rate of undocumented “self-transfer” among newer clients or a greater risk of disengagement.

Our study had several limitations that call for caution in interpretation of results, many of which are alluded to above. Data sources were incomplete and inaccurate, and the rates of tracing contact and return to care reported here reflect only our best estimates from available data; we are aware that data quality may bias our findings in either direction. The retrospective, observational design does not permit causal inference about whether tracing itself drives return to care, nor can we know if other interventions influenced return rates at the study sites differentially. Our sample was drawn from 15 facilities in three districts, and findings may not generalize to other provinces, tracing models, or countries. Many of those who never returned to the study sites are likely to have self-transferred to other facilities and resumed care[24]. We could not estimate the optimal timing of tracing following a missed visit or accurately estimate the overlap between telephone and physical tracing efforts. Finally, because of gaps in register availability (Figure 2), we could not determine what proportion of all tracing-eligible clients at each site were captured in our sample, which may introduce selection bias if register completeness is correlated with tracing effort or outcomes.

## Conclusions

The findings reported above suggest that tracing HIV treatment clients who have missed clinic visits or medication pickups in South Africa is associated with improvements in outcomes but, in its current form, also results in substantial wasted resources and may not affect client behavior as much as intended. Although missed appointments remain an important signal for identifying clients who may require follow-up, they should not automatically be interpreted as confirmed treatment interruptions or disengagement from care. The observed frequency of spontaneous return, together with instances of inaccurate tracing eligibility, suggests that tracing systems could benefit from more timely reconciliation of clinical attendance, medication collection, and client outcome information. Better data systems and standardization of documentation may help to distinguish missed appointments from confirmed treatment interruptions and disengagement from care, thus allowing better targeting of tracing efforts.

Based on these findings, we propose three programmatic recommendations. First, digitizing the paper tracing register into an electronic module directly linked to the EMR would ensure that tracing attempts, methods, and outcomes are automatically updated, reducing the number of misclassified clients and improving both documentation and the ability to measure tracing effectiveness at facility level. Second, our finding that a substantial number of clients who cannot be reached by tracing return to care on their own points to a need for more targeted tracing that focuses limited resources on clients who genuinely require outreach. Timing is important: early tracing likely expends resources on clients who will return on their own, while delayed tracing likely decreases the probability of both successful contact and return to care. Further research on optimal timing of tracing is critical. And third, interventions that promote self-initiated updates by clients and two-way communication, such as a mobile phone platform allowing clients to update contact details, confirm appointments, reschedule, or report a transfer themselves, could allow better allocation of tracing resources to those truly in need.

Even if tracing only modestly increases the overall proportion of clients who return to care, it may generate other benefits that should be considered in deciding how much to invest in it. Clients who were reached returned sooner than those who were not reached, suggesting that it may help avoid the harms of longer treatment interruptions and thus longer periods of viral non-suppression. Tracing also helps treatment programs determine the true status of those who disengage from care, by documenting mortality, transfer, and other final outcomes, improving estimates of retention and disengagement overall. From a programme perspective, our results thus support strengthening how tracing is conducted and documented, while retaining it as an integral component of the national treatment program.

## Supplementary files

Supplementary file 1: SOP 7

Supplementary figures 1-4: Flow charts showing sample construction, by district

Supplementary table 1: Client profile by tracing outcome

## Authors’ contributions

Conceptualization: NM, VN, SR

Funding acquisition: SR

Project administration: NM, VN

Supervision: MM1, LS, SR

Data collection and curation: VN, NM

Data analysis: NM

Writing – original draft: NM, SR

Writing – review, editing & revision: NM, VN, EK, SL, MM1, MB, MM2, LM, ARM, & SR

Final manuscript approval: NM, VN, EK, SL, MM, MB, MM, LM & SR

## Data availability

Tracing registers and TIER.Net data are owned by the South African National Department of Health. The TIER.Net data can be requested from Mr. Mbulelo Cabuko, Director, Health Information Research Monitoring and Evaluation (HIRME) Cluster, National Department of Health, Pretoria, South Africa.

## Supplementary Figures 1-4. Numbers of clients enrolled in the study, by district and tracing type

**Figure 1.**
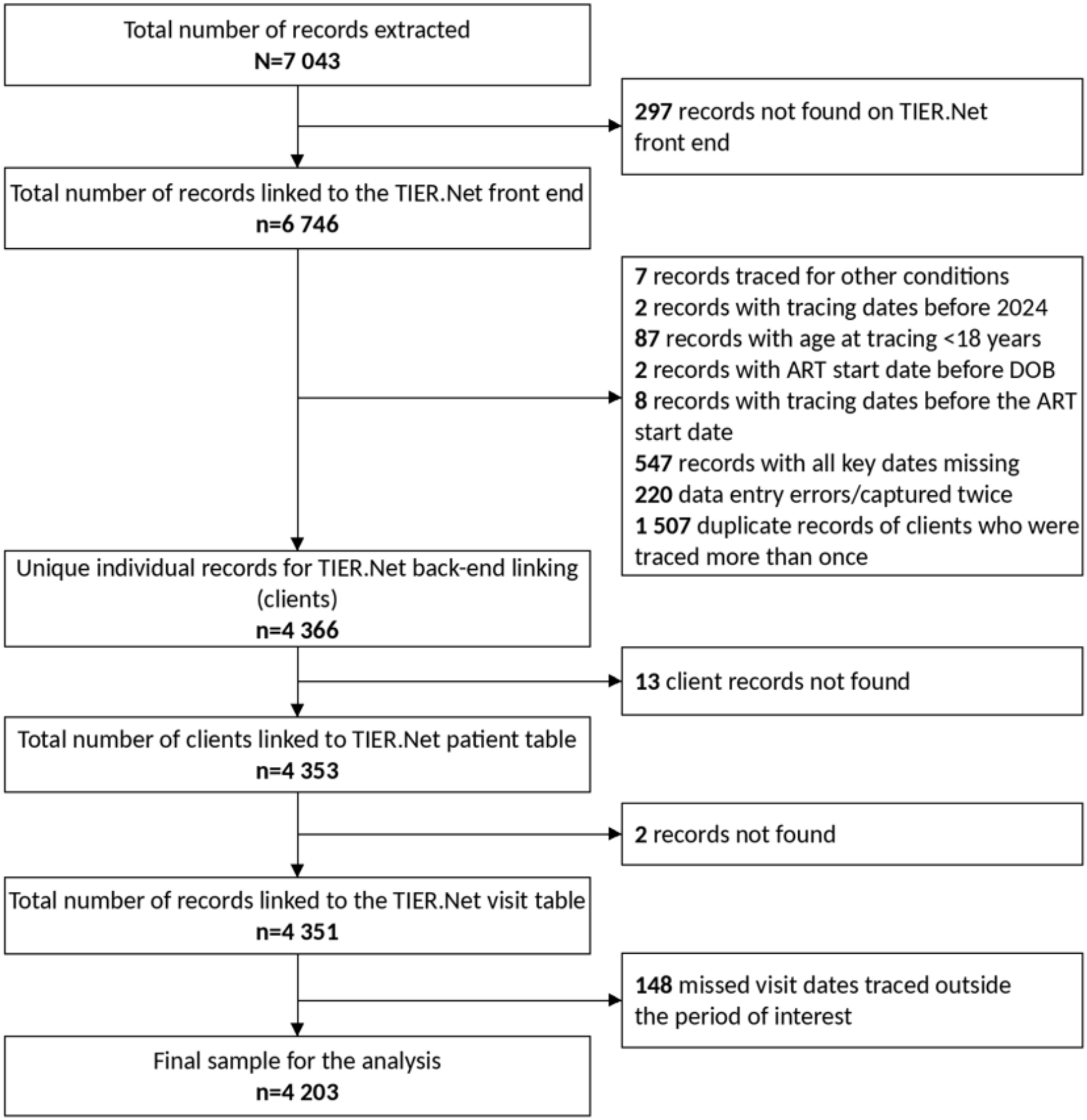
Telephone tracing – King Cetshwayo District

**Figure 2.**
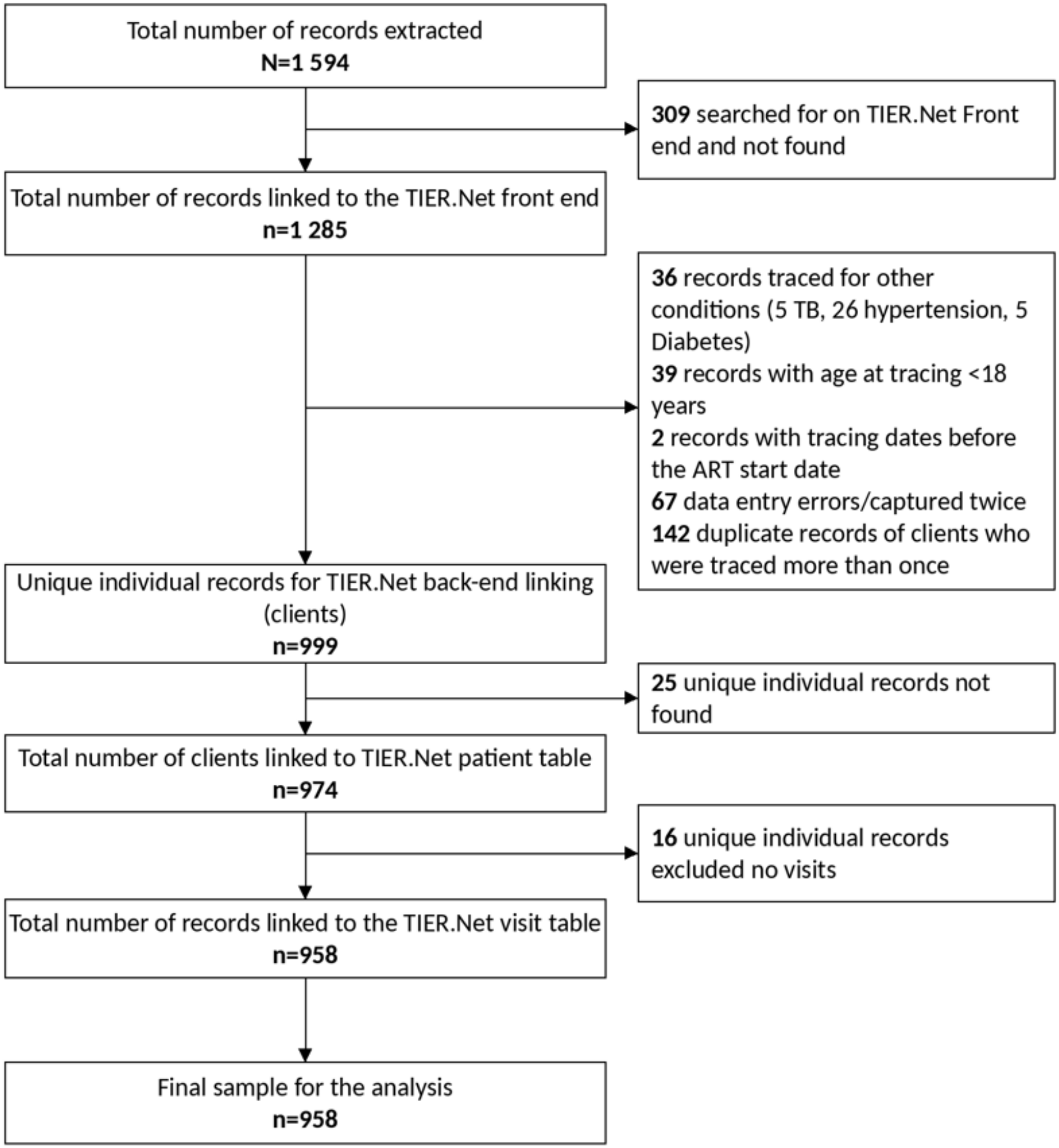
Telephone tracing – West Rand District

**Figure 3.**
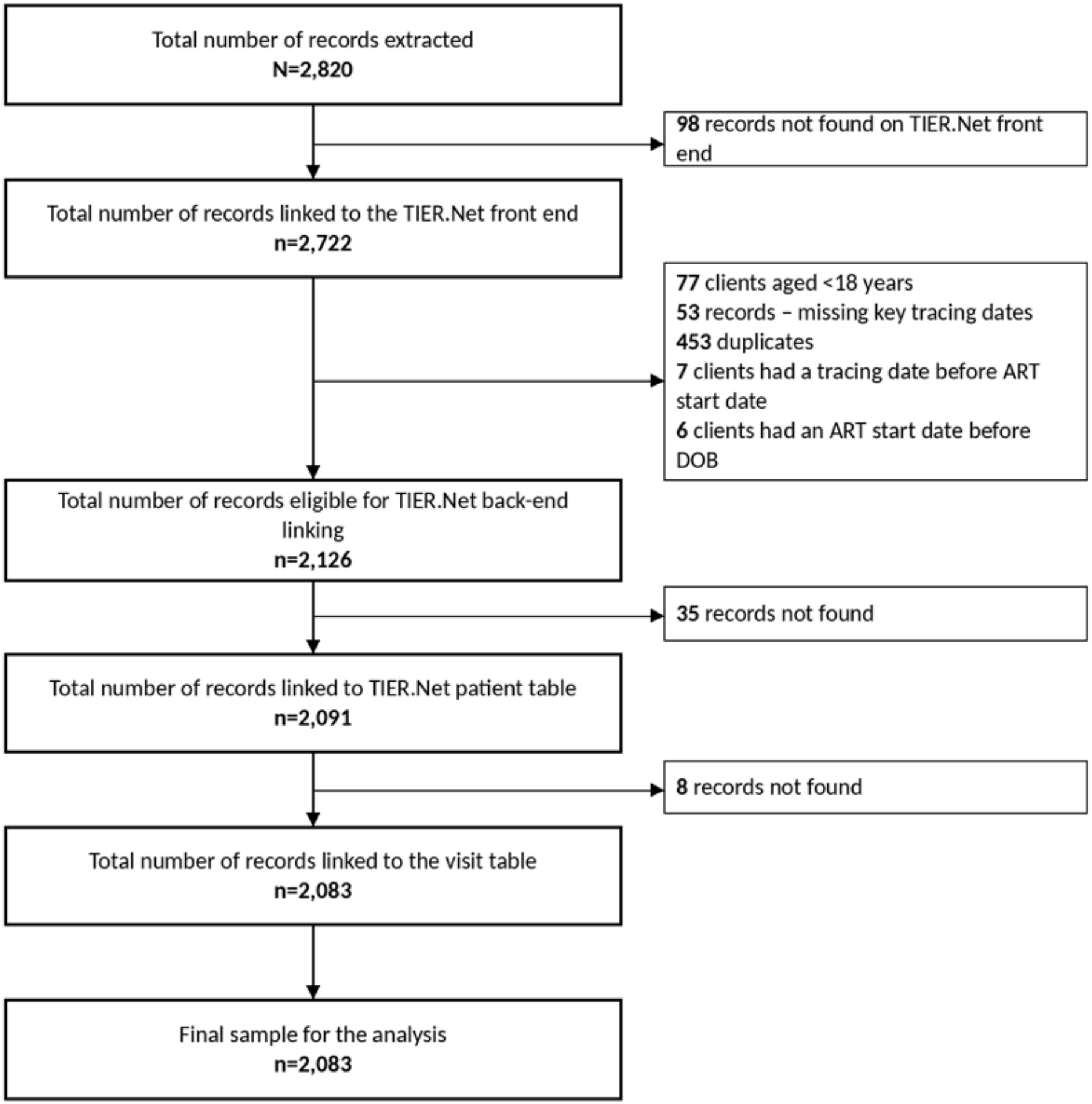
Physical tracing – Ehlanzeni District

**Figure 4.**
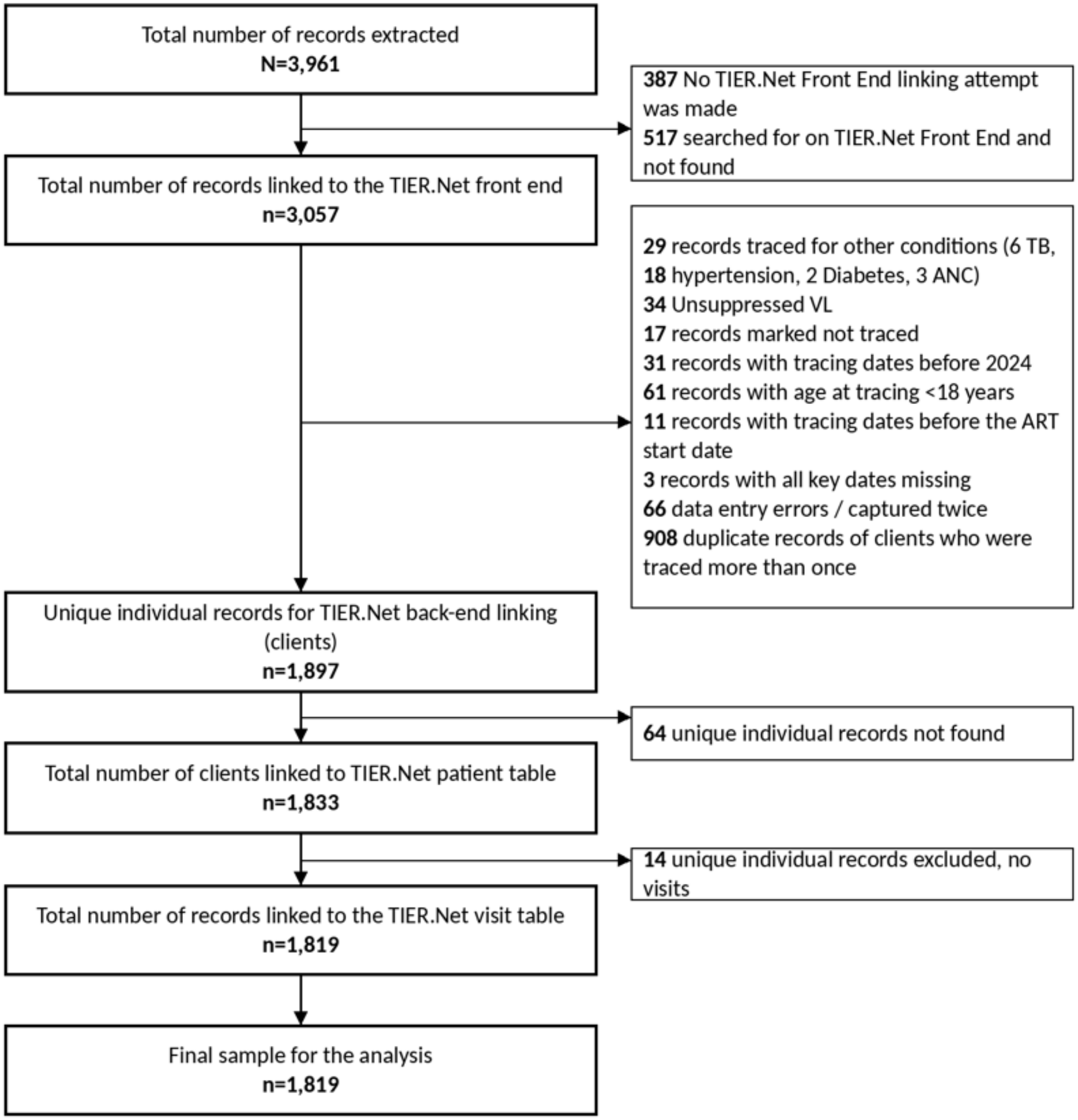
Physical tracing – West Rand District

**Supplementary Table 1.**
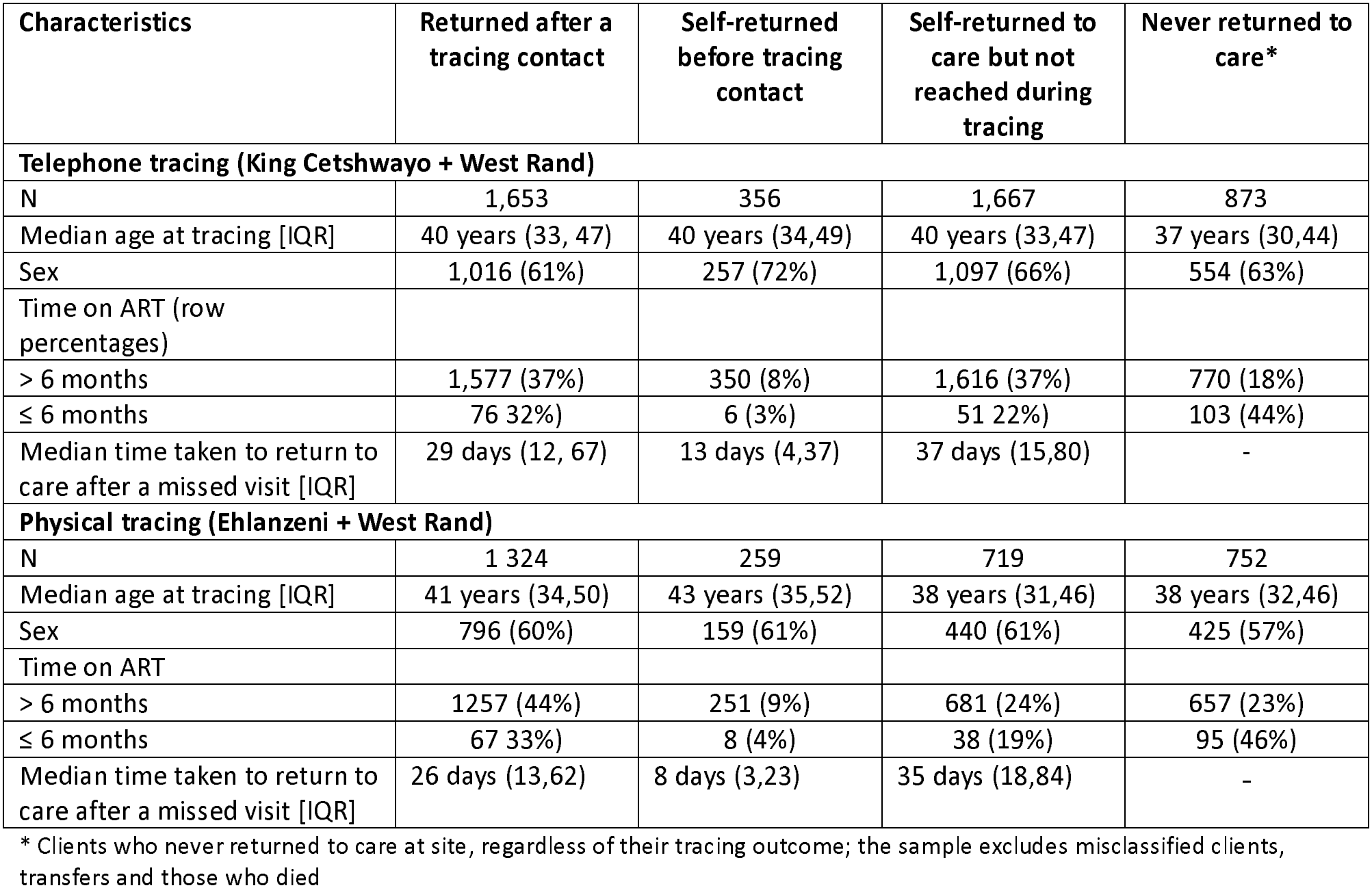
Client profile by tracing outcome.

| Characteristics | Returned after a tracing contact | Self-returned before tracing contact | Self-returned to care but not reached during tracing | Never returned to care* |
|---|---|---|---|---|
| **Telephone tracing (King Cetshwayo + West Rand)** |  |  |  |  |
| N | 1,653 | 356 | 1,667 | 873 |
| Median age at tracing [IQR] | 40 years (33, 47) | 40 years (34,49) | 40 years (33,47) | 37 years (30,44) |
| Sex | 1,016 (61%) | 257 (72%) | 1,097 (66%) | 554 (63%) |
| Time on ART (row percentages) |  |  |  |  |
| > 6 months | 1,577 (37%) | 350 (8%) | 1,616 (37%) | 770 (18%) |
| ≤ 6 months | 76 32%) | 6 (3%) | 51 22%) | 103 (44%) |
| Median time taken to return to care after a missed visit [IQR] | 29 days (12, 67) | 13 days (4,37) | 37 days (15,80) | - |
| **Physical tracing (Ehlanzeni + West Rand)** |  |  |  |  |
| N | 1 324 | 259 | 719 | 752 |
| Median age at tracing [IQR] | 41 years (34,50) | 43 years (35,52) | 38 years (31,46) | 38 years (32,46) |
| Sex | 796 (60%) | 159 (61%) | 440 (61%) | 425 (57%) |
| Time on ART |  |  |  |  |
| > 6 months | 1257 (44%) | 251 (9%) | 681 (24%) | 657 (23%) |
| ≤ 6 months | 67 33%) | 8 (4%) | 38 (19%) | 95 (46%) |
| Median time taken to return to care after a missed visit [IQR] | 26 days (13,62) | 8 days (3,23) | 35 days (18,84) | - |

\* Clients who never returned to care at site, regardless of their tracing outcome; the sample excludes misclassified clients, transfers and those who died

